# Biophysical modeling of corticospinal tract activation predicts motor contractions in subthalamic deep brain stimulation

**DOI:** 10.64898/2026.05.08.26352709

**Authors:** J Roediger, K Butenko, AP Krämer, IA Sahin, JK Behnke, S Oxenford, J Schikora, M Perales, MS Tuncer, T Picht, B Al-Fatly, GH Schneider, TA Dembek, AA Kühn

**Author notes:** **Corresponding author:** Jan Roediger, MD/PhD, Charité Universitätsmedizin Berlin, Department of Neurology, Movement Disorders and Neuromodulation Unit, Campus Mitte, Charitéplatz 1, Berlin 10117, Germany.

## Abstract

Stimulation-induced motor contractions are among the most common dose-limiting side effects in subthalamic nucleus deep brain stimulation for Parkinson’s disease, yet no quantitative models exist to predict their occurrence from imaging data. Here, we combine pathway activation modeling of the corticospinal and corticobulbar tracts within the posterior limb of the internal capsule with a data-driven prediction framework. Evaluated by leave-one-patient-out cross-validation across an intraoperative (42 patients, 352 sites) and postoperative sub-cohort (11 patients, 176 contacts), the model explained 31-35% of the variance in observed motor contraction thresholds. The model reliably identified contacts with the lowest and highest motor contraction thresholds within individual electrodes, with strongest performance for distinguishing directional segments at the same electrode level (84% and 64% accuracy; p < 0.001), supporting its potential value for postoperative DBS programming. Back-projection of model coefficients provided an anatomically interpretable mapping consistent with known capsular anatomy. The framework is openly available and may inform computational tools for surgical planning, intraoperative validation, and postoperative programming in DBS.

## Introduction

Deep brain stimulation (DBS) is an established neurosurgical treatment for movement disorders, with over 200,000 devices implanted worldwide.^1,2^ Electrodes are implanted in subcortical brain structures to deliver electrical stimulation that modulates pathological neural circuit activity. While highly effective, the clinical outcome of DBS critically depends on precise electrode placement and careful selection of stimulation parameters.^3^ Both are guided by the aim to maximize modulation of therapeutic target structures while avoiding co-stimulation of adjacent fiber tracts, which can induce side effects.

Corticospinal and corticobulbar fiber tracts (CSBT) within the posterior limb of the internal capsule (PLIC) lie in close anatomical proximity to the subthalamic nucleus (STN), the most common DBS target for Parkinson’s disease.^4^ Their co-stimulation during DBS leads to tonic muscle contractions contralateral to the stimulated hemisphere.^5^ The motor contraction threshold, the stimulation amplitude at which these contractions first occur, is determined by the spatial relationship between the stimulation site and the CSBT and represents a hard upper limit on the stimulation amplitude that can be applied. Ensuring a sufficiently high motor contraction threshold is therefore addressed at multiple stages of DBS treatment, from surgical trajectory planning and intraoperative test stimulations to postoperative contact selection. ^6,7^ Quantitative tools to predict this threshold could support clinical decision-making at each stage.

Over the past two decades, computational models of DBS-induced neural activation have been developed. These models combine simulations of the electric field generated by the DBS electrode based on the finite element method (FEM) with multi-compartment cable models of myelinated axons to estimate whether individual axons are activated at a given stimulation amplitude.^8,9^ Applied to anatomically reconstructed fiber pathways, this pathway activation modeling (PAM) framework enables estimation of which neural structures are recruited for a given electrode position and stimulation setting.^10,11^ Stimulation-induced motor contractions provide a uniquely suitable testbed for these models since the underlying mechanism is well characterized and the clinical readout is immediate, unambiguous, and reproducible. Early studies leveraged these properties to calibrate biophysical models against observed motor thresholds in individual patients.^12,13^ More recent studies have shown that detailed biophysical CSBT models outperform simplified approaches in predicting electrophysiological surrogate markers of tract activation.^14,15^ Yet the direct link to the clinical outcome of stimulation-induced motor contractions has not been established.

In parallel, data-driven approaches have leveraged the anatomical variability of DBS electrode placement across patients to build quantitative prediction models across various symptoms and indications. ^16,17^ By linking electrode location and stimulation parameters to clinical outcomes at the population level, these models pave the way for empirically grounded, automated DBS programming algorithms.^18-20^ However, existing approaches have focused primarily on therapeutic effects, while stimulation-induced motor contractions have not been addressed despite the critical role of side-effect avoidance in clinical programming. Furthermore, the biophysical features used in data-driven DBS models are typically limited to simplified surrogates such as volumes of tissue activated or electric field estimates, since the anatomical and biophysical foundations of therapeutic DBS effects remain incompletely understood. The CSBT provides an opportunity to move beyond these surrogates, as its macroscopic organization^21^ and microscopic axon properties^22^ have been characterized in detail, enabling biophysically grounded feature engineering.

Building on these foundations, we develop a hybrid framework combining biophysically detailed pathway activation modeling of the CSBT with a data-driven prediction model for stimulation-induced motor contractions in STN-DBS (*Fig. 1*). The framework exemplifies a physics-informed machine learning approach: biophysical simulations provide physically meaningful features that are calibrated against clinical outcomes, bridging mechanistic modeling and patient-specific prediction. The model is developed on an intraoperative cohort and a postoperative sub-cohort and evaluated using leave-one-patient-out cross-validation. Beyond population-level performance, we additionally assess the model’s ability to identify contacts with the highest and lowest motor contraction thresholds at the individual electrode level, providing a direct measure of its potential utility for postoperative DBS programming. Back-projection of model coefficients onto the underlying tract anatomy provides an interpretable spatial mapping of predicted motor contraction risk. To facilitate adoption, the model and code are publicly available with the aim of providing computational tools that may inform clinical decision-making from surgical planning to postoperative programming.

**Figure 1.**
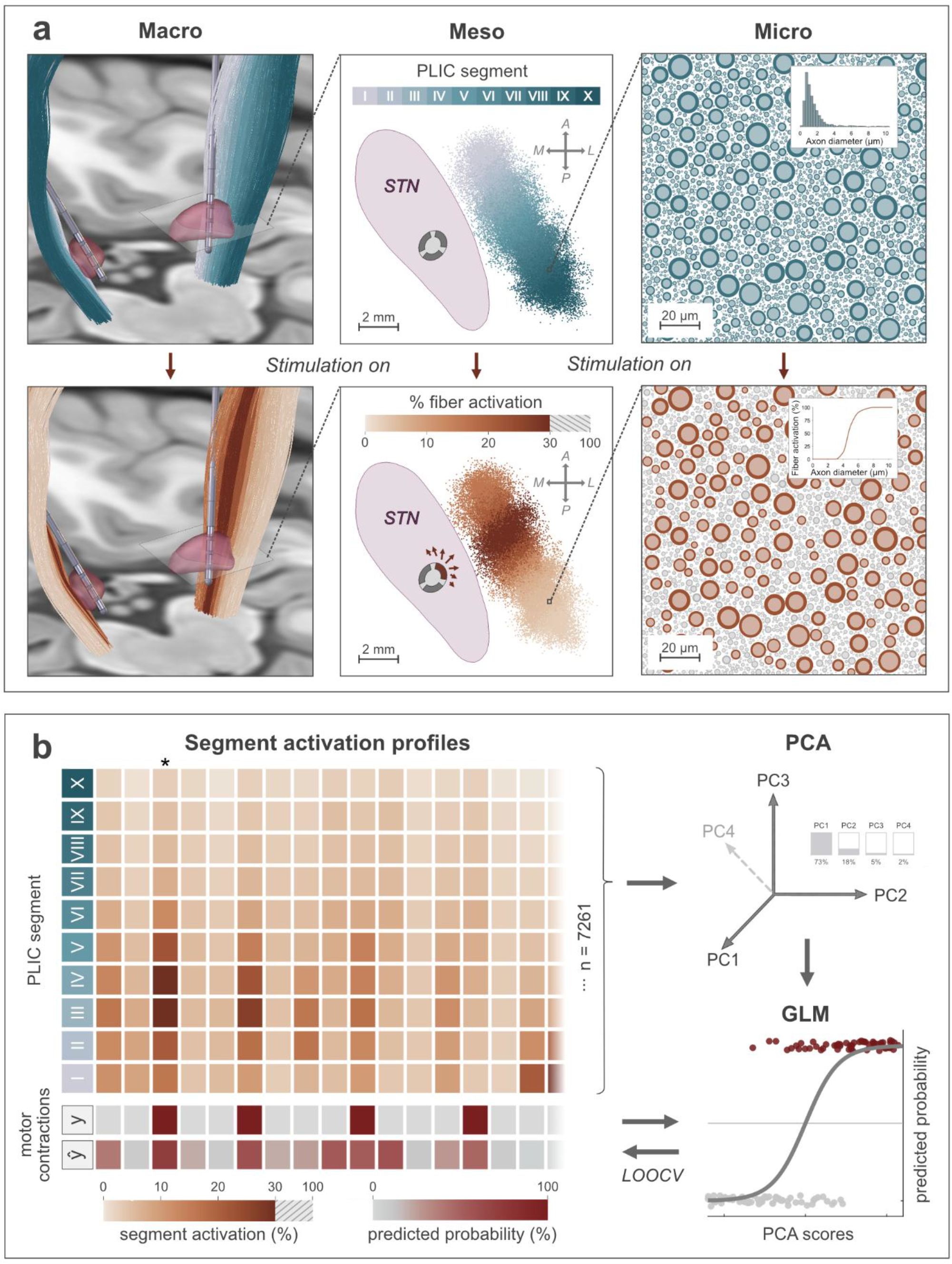
Biophysical modeling and statistical prediction framework for stimulation-induced motor contractions. (a) Multiscale biophysical modeling of corticospinal tract activation. Pathway activation modeling was performed across three anatomical scales, shown here for one example stimulation site. The upper row illustrates the anatomical and biophysical setup, the lower row the corresponding state under active stimulation. *Upper row - Model setup. Macro:* DBS electrodes were localized following the pipeline described in Section 4.4. The normative posterior limb of the internal capsule (PLIC) was warped from MNI space into individual patient space using patient-specific nonlinear deformation fields. *Meso:* Shown is an axial cross-section through the PLIC at STN level. The normative PLIC had been subdivided into ten segments along the anteromedial-to-posterolateral axis (segments I-X) to capture the spatial heterogeneity of corticospinal fiber organization at this level. *Micro:* Fibers within the PLIC span a broad range of axon diameters. Their distribution was approximated from electron microscopic cross-sections of the non-human primate corticospinal tract (inset histogram showing distribution taken from Firmin et al.). *Lower row - Active stimulation. Macro & Meso:* with stimulation switched on, the color gradient (white to brown) encodes the proportion of activated fibers per segment, both along the streamlines and in the axial cross-section. *Micro:* whether an individual fiber is activated by the extracellular electric field was determined using multi-compartment cable models of myelinated axons. Because activation thresholds depend strongly on axon diameter (inset), the % fiber activation per segment shown in the Macro and Meso panels was computed in accordance with the empirical diameter distribution. **(b) Statistical prediction pipeline**. For each stimulation site and amplitude step, the ten-dimensional segment activation profile (left, rows: PLIC segments I-X; columns: individual observations; n = 7261 across both cohorts) together with the observed binary motor contraction outcome (y) entered the statistical model. The column marked with the asterisk corresponds to the activation profile of the example observation visualized in the lower row of Panel A. Principal component analysis (PCA) was applied for dimensionality reduction and to address multicollinearity between segment activation profiles. The retained PCA scores, together with stimulation amplitude and a condition-specific interaction term (intraoperative vs. postoperative), entered a generalized linear model (GLM) with a logistic link function to predict motor contraction probability (ŷ). Model performance was evaluated by leave-one-patient-out cross-validation (LOOCV).

## Results

### 2.1 Cohort Characteristics

A total of 42 patients with Parkinson’s disease (79 hemispheres, 352 stimulation sites) fulfilled inclusion criteria for the intraoperative cohort, of whom 11 (22 hemispheres, 176 contacts) additionally completed postoperative assessments and constituted the postoperative sub-cohort. A trial profile is provided in the *Suppl. Fig. 1* and demographic and clinical characteristics for both study phases are summarized in *Table* 1. The spatial distribution of all tested stimulation locations is shown in Suppl. Fig. 6.

**Table 1.**
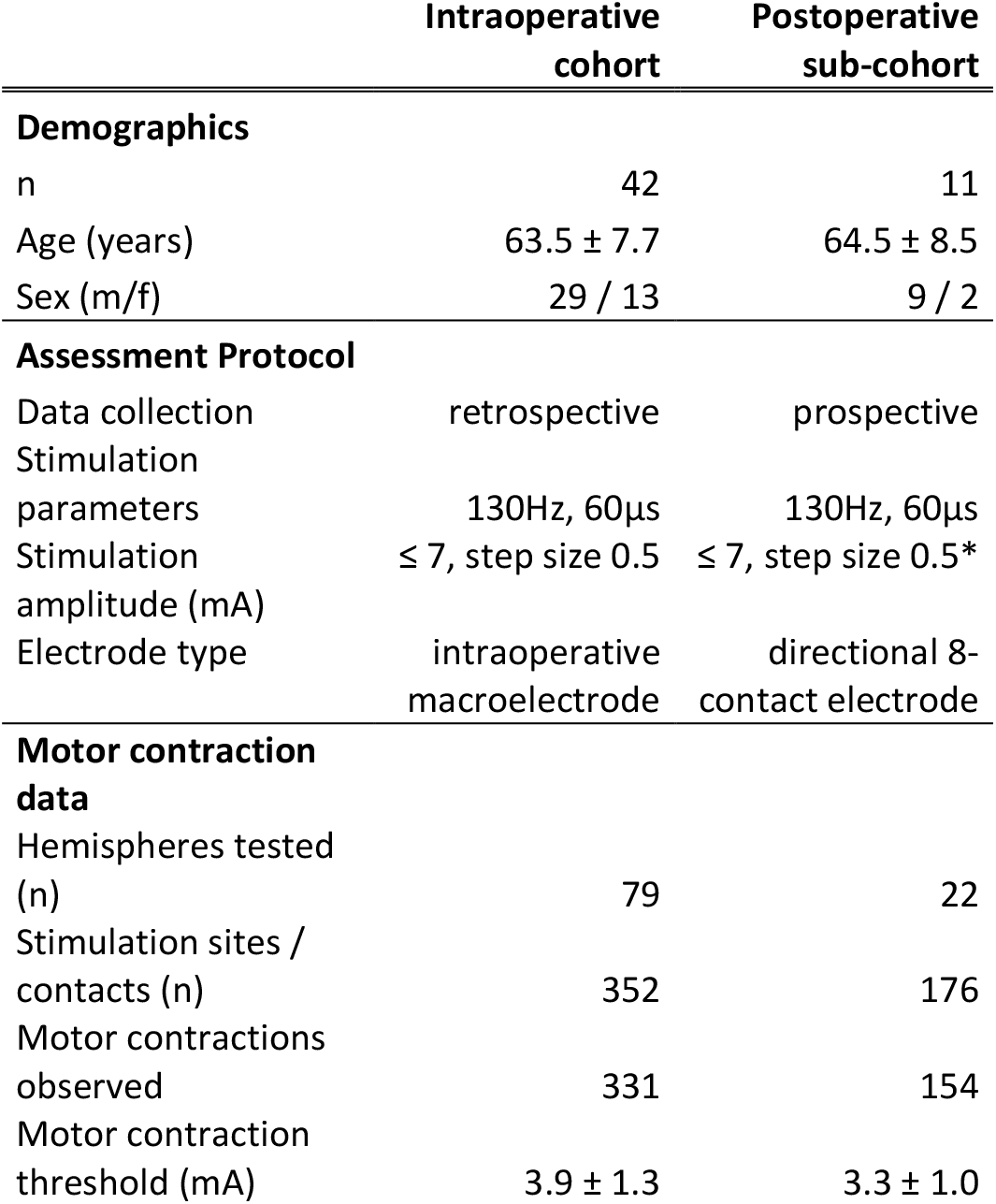
Cohort characteristics and motor contraction assessment. Demographic, procedural, and descriptive data for the intraoperative cohort and the postoperative sub-cohort. Age and motor contraction threshold are reported as mean ± standard deviation. Stimulation was delivered in monopolar cathodal configuration in both study phases with stimulation amplitudes increasing up to 7mA or until motor contractions or intolerable side-effects occurred. *Upon detection of motor contractions in the postoperative sub-cohort, the threshold was refined in 0.1 mA steps (see Section 4.3).

|  | Intraoperative cohort | Postoperative sub-cohort |
| --- | --- | --- |
| **Demographics** |  |  |
| n | 42 | 11 |
| Age (years) | $63.5 \pm 7.7$ | $64.5 \pm 8.5$ |
| Sex (m/f) | 29 / 13 | 9 / 2 |
| **Assessment Protocol** |  |  |
| Data collection | retrospective | prospective |
| Stimulation parameters | 130Hz, 60μs | 130Hz, 60μs |
| Stimulation amplitude (mA) | $\leq 7$, step size 0.5 | $\leq 7$, step size 0.5* |
| Electrode type | intraoperative macroelectrode | directional 8-contact electrode |
| **Motor contraction data** |  |  |
| Hemispheres tested (n) | 79 | 22 |
| Stimulation sites / contacts (n) | 352 | 176 |
| Motor contractions observed | 331 | 154 |
| Motor contraction threshold (mA) | $3.9 \pm 1.3$ | $3.3 \pm 1.0$ |

### 2.2 Input Data and Internal Consistency

Motor contractions were elicited at 331 of 352 intraoperative stimulation sites (94%) and 154 of 176 postoperative contacts (88%), with mean clinical motor thresholds of 3.9 ± 1.3 mA and 3.3 ± 1.0 mA, respectively. Correlation between clinical and EMG-derived thresholds was strong across both assessment phases (intraoperative: r = 0.82; postoperative: r = 0.89; p < 0.001), with mean EMG thresholds lower than clinical by 0.37 mA intraoperatively and 0.26 mA postoperatively (*Suppl. Fig. 2*).

Each stimulation site was tested in discrete 0.5 mA steps up to a maximum of 7 mA or until discontinuation due to intolerable side effects. Encoding each amplitude step as a binary outcome (motor contraction present or absent, with all suprathreshold amplitudes classified as contraction present; *see Section 4.7*) yielded a total of 7,261 binary observations (4,928 intraoperative, 2,333 postoperative), of which 3,457 (48%) were classified as motor contraction present and 3,804 (52%) as absent.

The internal consistency of the two electrode localization approaches (*see Section 4.4*) was assessed by comparing intraoperative stereotactic coordinate-based and postoperative CT-based reconstructions across all patients. The median Euclidean distance between corresponding electrode locations was 1.08 mm (IQR 0.74-1.45 mm; range 0.22-3.93 mm; *Suppl. Fig. 3*).

### 2.3 Classification performance

Principal component analysis (PCA) was applied to the PLIC activation profiles across all stimulation sites and amplitude steps from both cohorts *(Fig. 1b)*. Four principal components were retained based on cross-validated threshold prediction performance (*Suppl. Fig. 4*), together explaining 98% of the total variance in segment-level activation. These four components, together with stimulation amplitude and the condition-specific interaction terms (see *Section 4.7*), entered the Generalized linear model (GLM). In the full model estimated on all available data, all terms reached statistical significance at p < 0.001 (*Suppl. Table 1*).

Model performance was evaluated using leave-one-patient-out cross-validation. The model achieved an AUC of 0.95 and 0.97 for the intra- and postoperative cohorts, respectively *(Fig. 2a, c)*. To isolate the contribution of PLIC-component activation from the effect of stimulation amplitude, classification performance was additionally evaluated at fixed amplitude levels within the clinically relevant range (2-5 mA). Classification performance remained strong with AUC-values between 0.70 and 0.86 within the intraoperative and between 0.78 and 0.93 within the postoperative sub-cohort, confirming that PLIC segment activation profiles carry discriminative information across clinically relevant amplitude ranges.

**Figure 2.**
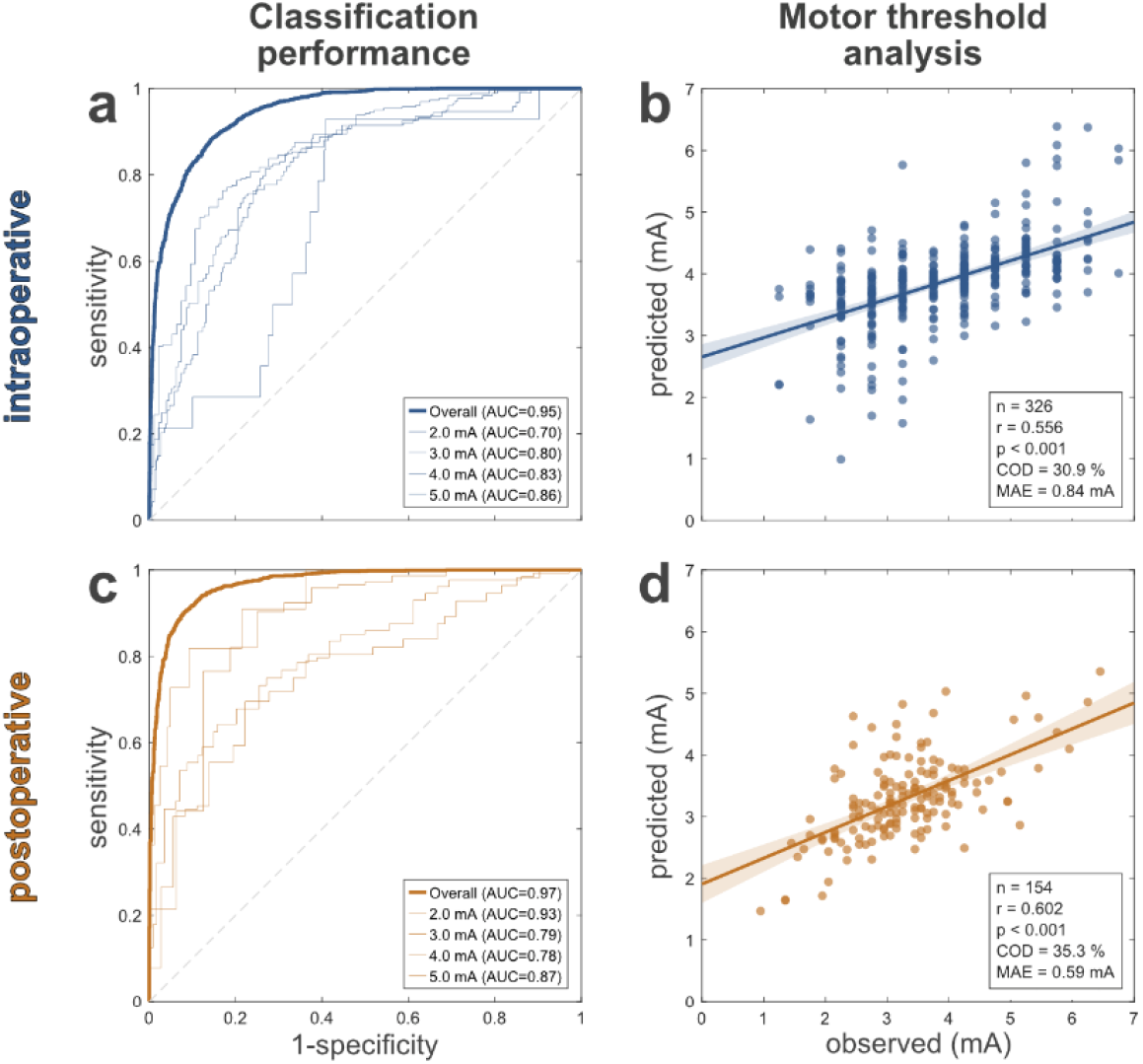
Cross-validated model performance across intraoperative cohort and postoperative sub-cohort. Intraoperative results are shown in blue (top row), postoperative results in orange (bottom row). All analyses were performed using leave-one-patient-out cross-validation. **(a, c) Classification performance**. Receiver operating characteristic curves for the classification of stimulation-induced motor contractions in the intra- (a) and postoperative (c) cohorts. Bold curves show overall classification performance across all amplitude steps; thin curves show performance at fixed amplitude levels (2-5 mA). Area under the curve (AUC) values are reported in the legends. The dashed diagonal indicates chance-level performance. **(b, d) Motor threshold analysis**. Predicted versus observed motor contraction thresholds for the intraoperative cohort (b) and postoperative sub-cohort (d). Each point represents one stimulation site. Predicted thresholds were derived by linear interpolation of the estimated probability curve to identify the amplitude at which a 50% contraction probability was predicted. Solid lines denote the linear regression fit with 95% confidence bands. Inset statistics report sample size (n), Pearson correlation coefficient (r), coefficient of determination (COD), and mean absolute error (MAE).

### 2.4 Motor Threshold Analysis

Predicted motor contraction thresholds (see *Section 4.7*) were compared against empirically observed clinical thresholds across both cohorts. The model explained 30.9% of the variance (COD) in intraoperative thresholds (r = 0.56, MAE = 0.84 mA, n = 326, p < 0.001; *Fig. 2d*) and 35.3% in the postoperative sub-cohort (r = 0.60, MAE = 0.59 mA, n = 154, p < 0.001; *Fig. 2b*).

### 2.5 Contact-Level Predictions

The implanted directional DBS leads (Medtronic SenSight×) comprise eight contacts across four levels (*Fig. 3b*). A clinically relevant application of the model is to identify which contact to preferentially activate or avoid to prevent motor contractions. To this end, for each of the 22 postoperative directional DBS leads, the contacts with the highest and lowest predicted motor thresholds were identified and compared against empirical observations *(Fig 3)*.

**Figure 3.**
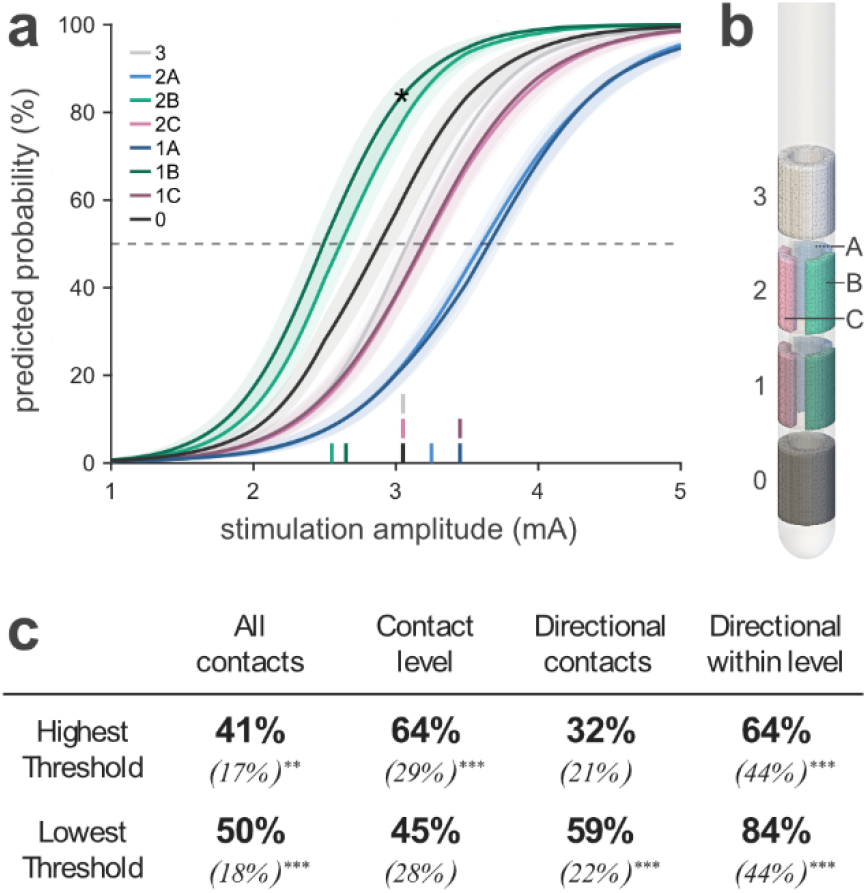
Within-electrode identification of contacts with lowest and highest motor contraction thresholds in the postoperative sub-cohort. **(a)** Predicted motor contraction probability as a function of stimulation amplitude for each contact of a representative directional DBS lead. Shaded bands denote 95% confidence intervals; the dashed horizontal line marks the 50% probability criterion used to define the predicted motor threshold. Rug marks along the x-axis indicate amplitudes at which motor contractions were clinically observed. The asterisk marks the example observation also shown in Figure 1 (lower row of Panel a and highlighted column in Panel b). **(b)** Schematic of the 8-contact directional DBS lead (Medtronic SenSight™): a bottom ring contact (level 0), two directional levels with three segments each (levels 1 and 2; segments A, B, C), and a top ring contact (level 3). Contact colors correspond to the curves in Panel A. **(c)** Proportion of leads (n = 22) for which the model correctly identified the contact with the lowest or highest empirically observed motor threshold, evaluated under four grouping schemes: *All contacts*: Ranking across all eight contacts; *Contact level*: Contacts grouped into the four levels, with directional segments within a level pooled; *Directional contacts*: Ranking restricted to the six directional contacts (1A-2C); *Directional within level*: Ranking restricted to the three segments within the same directional level (1A/1B/1C and 2A/2B/2C, evaluated separately; n = 44). Values in parentheses denote chance-level performance. Asterisks indicate significance against chance: \**p* < 0.05, \*\**p* < 0.01, \*\*\**p* < 0.001.

The model correctly identified the contact with the lowest motor threshold in 50% of leads when considering all eight contacts (chance level: 18%, p < 0.001), and in 59% when restricted to the six directional contacts (chance: 22%, p < 0.001). Correct identification of the electrode level with the lowest threshold was achieved in 45% of leads (chance: 28%, p = 0.055), whereas identification of the lowest-threshold directional contact within the same electrode level was substantially higher at 84% (chance: 44%, p < 0.001). Results for identification of the contact with the highest motor threshold are shown in *Fig. 3c*.

### 2.6. Sensitivity analysis

Re-estimating the model using EMG-derived instead of clinically observed motor thresholds yielded consistent classification (AUC: 0.93 intraoperative, 0.96 postoperative) and threshold prediction performance (COD: 24.6% intraoperative, 32.8% postoperative; both p < 0.001). Estimating separate models on the intraoperative cohort and postoperative sub-cohort independently also yielded comparable results (intraoperative: AUC 0.95, COD 30.8%; postoperative: AUC 0.97, COD 39.1%; both p < 0.001). Dysarthria, which was reported at 55 of 176 contacts (31.3%) within the postoperative sub-cohort, was included alongside motor contractions as an extended endpoint in an additional sensitivity analysis, yielding comparable performance (AUC 0.97, COD 32.6%). A comprehensive comparison of all sensitivity analyses is provided in *Suppl. Table 2*.

### 2.7 Anatomical Mapping

Back-projection of GLM coefficients onto the ten PLIC segments revealed a gradient along the anteromedial-to-posterolateral axis, with posterolateral segments carrying the largest predictive weights *(Fig. 4a)*. Coefficients peaked at segments VIII and IX (0.21 and 0.23, respectively), with a moderate decline at the most posterolateral segment X (0.14); all three differed significantly from zero. Anteromedial segments I-VI exhibited near-zero weights with confidence intervals spanning or approaching zero, indicating a negligible contribution to predicted motor contraction probability.

**Figure 4.**
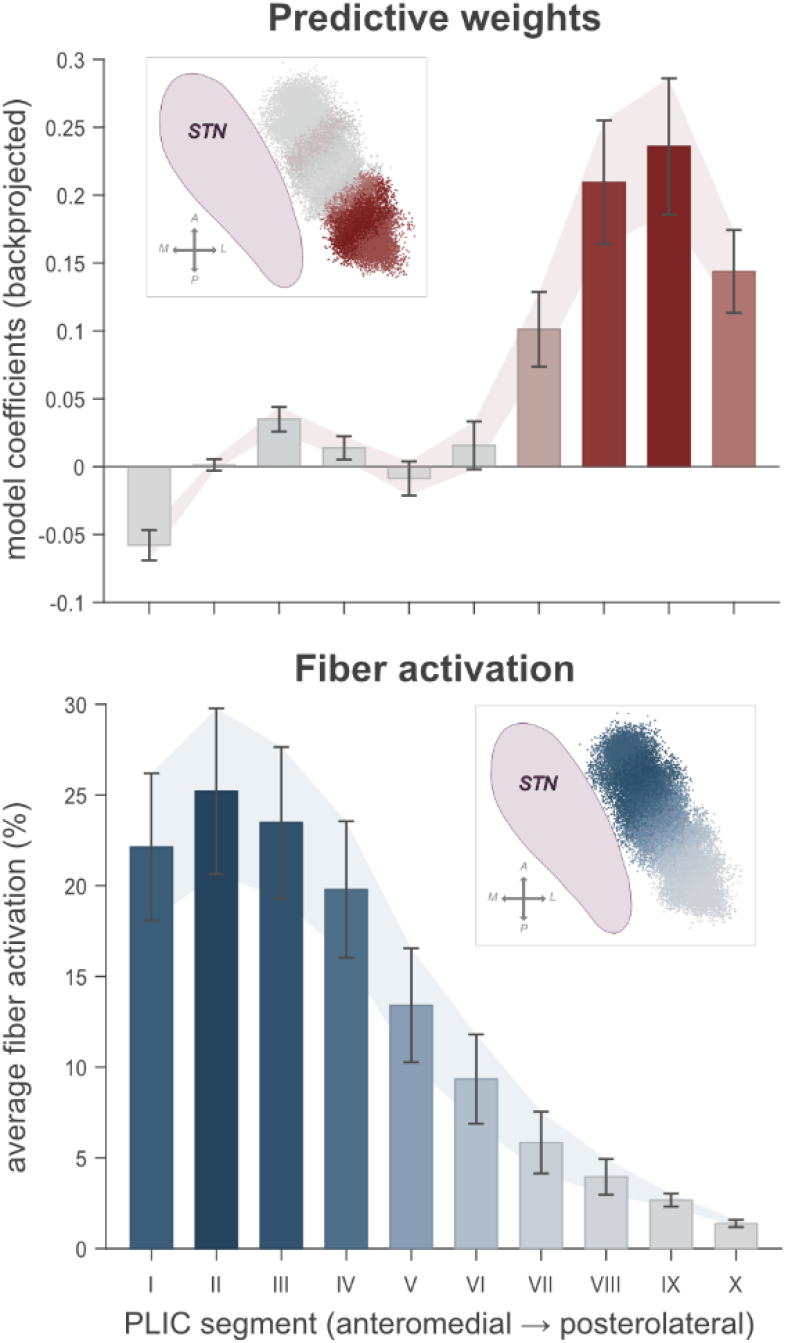
Anatomical mapping of model coefficients and fiber activation across PLIC segments. Segments are ordered from anteromedial (I) to posterolateral (X). Results are shown for the postoperative sub-cohort; comparable results for the intraoperative cohort are provided in Suppl. Fig. 5. **(a)** Back-projected GLM coefficients for each PLIC segment, reflecting the relative contribution to predicted motor contraction probability (logit scale). Positive values indicate that activation of a given segment increases the predicted probability of motor contractions. Coefficients were obtained by projecting GLM regression weights through the PCA loading matrix onto the original ten segments. Error bars denote 95% confidence intervals. **(b)** Mean fiber activation (%) per PLIC segment at the cohort-specific mean observed motor threshold amplitude (3.3 mA). Error bars denote 95% confidence intervals of the mean. The contrasting gradients across panels illustrate that the segments most predictive of motor contractions (posterolateral, Panel A) are those least activated at the mean observed motor threshold amplitude (Panel b). STN: subthalamic nucleus; A: anterior, P: posterior, M: medial, L: lateral.

The average activation profile at the mean observed motor threshold of 3.3 mA within the postoperative cohort revealed a contrasting spatial pattern: fiber recruitment was concentrated in the anteromedial PLIC, peaking at 25% in segment II and declining to 1% in posterolateral segment X *(Fig. 4b)*. Thus, the segments most predictive of motor contractions were those least activated at the mean observed motor threshold. Results are shown for the postoperative cohort; comparable results for the intraoperative cohort are provided in *Suppl. Fig. 5*.

## Discussion

This study demonstrates that stimulation-induced motor contractions in STN-DBS can be estimated probabilistically by combining biophysical pathway activation modeling with a data-driven prediction framework. The model achieved strong classification and threshold prediction performance across both cohorts (AUC 0.95-0.97, COD 31-35%), differentiated contacts with highest and lowest motor thresholds on individual electrodes, and yielded an anatomically interpreg of motor contraction risk across the PLIC. These results suggest potential applications across the clinical workflow of DBS, from surgical planning and intraoperative validation to imaging-guided postoperative programming.

Our biophysical modeling pipeline builds on established computational DBS models that couple FEM-based electric field simulations with multi-compartment cable models along reconstructed fiber trajectories.^8-11^ While such forward models have been validated against electrophysiological markers for the CSBT,^14,23^ predicted activation levels cannot be directly translated into clinical effect probabilities without calibration. Conversely, data-driven approaches have enabled calibrated predictions of therapeutic outcomes^16,17^ but have not been applied to motor contractions. Our study bridges this gap, combining detailed biophysical modeling of the corticospinal tract with a data-driven prediction framework to predict stimulation-induced motor contractions. The CSBT is particularly amenable to this hybrid approach because its macroscopic organization^21^ and microscopic axon properties^22^ are well characterized, enabling biophysically grounded feature engineering at both scales. At the macroscopic level, the ten-segment parameterization allowed the statistical model to learn the spatial distribution of motor-relevant fibers from clinical data. At the microscopic level, incorporating the empirical diameter distribution replaced the commonly used single-diameter assumption with a biophysically realistic representation of axonal heterogeneity. The biophysical modeling pipeline thus transforms a high-dimensional problem into a compact set of physically meaningful features: spatially resolved, axon-diameter-weighted activation profiles across the PLIC. These features serve as input to a statistical model that calibrates them against observed clinical outcomes, forming a physics-informed prediction framework that translates biophysically grounded features into calibrated clinical predictions.

In such hybrid frameworks, not only the feature engineering but also the choice of statistical model should be guided by prior knowledge about the underlying mechanism. The relationship between CSBT fiber activation and motor contraction probability is expected to follow a monotonically increasing, sigmoidal pattern: at low activation levels, the probability of a clinically detectable contraction is negligible; beyond a certain threshold, contractions become highly likely. A GLM with a logistic link function captures this relationship directly. Moreover, for a model with potential clinical application, predictability and transparency are essential, and the GLM produces no unexpected nonlinear effects. Finally, the limited dataset size (42/11 patients) favors parsimonious models since more complex architectures would risk overfitting and may not generalize reliably.

PCA served to address the strong collinearity among adjacent PLIC segments, which are co-activated due to their spatial proximity within the electric field. The optimal model complexity was determined by forward selection of principal components based on cross-validated performance. This approach generalizes beyond the present application: in settings where the target tract is less clearly delineated and activation of multiple surrounding pathways must be considered^24^, the same framework could accommodate higher-dimensional feature spaces, with regularization determining the appropriate model complexity in a data-driven manner.

To ensure reliable model training and evaluation, data quality was internally validated at multiple levels: Clinical and EMG-derived thresholds agreed closely (r = 0.82-0.89), the two electrode localization approaches showed concordance within the voxel-resolution, and sensitivity analyses confirmed that the findings are not driven by a single cohort or assessment modality (*Suppl. Table 2*). The consistency of results across two clinical settings that differ in electrode type, localization method, and patient state suggests that the physically informed features capture mechanistically relevant aspects rather than setting-specific confounds, a recognized advantage of physics-informed over purely data-driven approaches.^25^

Despite the consistent performance across both cohorts, the model required condition-specific calibration: interaction terms between study condition (intra-vs postoperative) and biophysical features were statistically significant, indicating systematic differences not captured by the biophysical simulation. Factors such as fibrotic encapsulation of implanted electrodes,^26^ or local tissue heterogeneity may contribute. These effects could in principle be modeled biophysically but would introduce patient-specific parameters that are unknown a priori, potentially adding rather than reducing uncertainty. Population-level calibration offers a pragmatic alternative by absorbing residual effects empirically. More broadly, quantities not reliably known from first principles, such as the spatial weighting of CSBT regions, should be estimated from data rather than imposed as potentially incorrect biophysical constraints. The biophysical model thus provides physically grounded features that transfer across settings, while the statistical component calibrates them against clinical reality.^27^

Back-projection of GLM coefficients confirmed the expected posterolateral concentration of motor-relevant CSBT fibers within the PLIC.^21^ The anatomical dissociation between predictive weight (posterolateral) and actual recruitment at therapeutic amplitudes (anteromedial, *Fig. 4*) offers a structural explanation for the therapeutic window in STN-DBS: at typical amplitudes, stimulation recruits primarily anteromedial fibers that do not elicit contractions, while the posterolateral corticospinal fibers remain largely unrecruited. The ability to map model coefficients back onto defined anatomical regions illustrates a key advantage of physically meaningful features: the resulting spatial patterns are directly interpretable in anatomical and clinical terms. This aligns with connectomic and tract-based mapping approaches that leverage DBS as a causal probe to infer which neural structures mediate clinical effects.^28^ As with any such approach, however, predictive features do not necessarily correspond to the anatomical structures causally mediating the effect (see Limitations).^29^

While the anatomical mapping results should be interpreted with the above caveats, the model’s primary value lies in its predictive capacity, specifically its ability to estimate motor contraction probabilities across the amplitude range rather than threshold amplitudes alone. We envision three concrete applications:

### Surgical planning

During stereotactic planning, motor contraction risk is typically estimated from proximity to the PLIC on two-dimensional cross-sectional images. However, simple distance measures do not account for the spatial organization of fibers or the directional properties of the electric field, and correlate only moderately with observed motor thresholds.^5^ The present model could instead provide motor contraction probability curves for candidate electrode positions, allowing the surgical team to evaluate trajectories by their predicted motor contraction profile. In combination with models of therapeutic benefit, this could move surgical planning toward quantitative optimization of the predicted therapeutic window.

### Intraoperative validation

During DBS surgery, the model could be integrated into intraoperative visualization tools such as Lead-OR^30^ to provide quantitative predictions of expected motor thresholds at each test stimulation site. Currently, this assessment relies on clinical experience. Comparing predicted and observed thresholds in real time could objectify this process: for instance, thresholds consistently lower than predicted could indicate deviation toward the CSBT, prompting repositioning before permanent implantation.

### Postoperative programming

In clinical practice side-effect thresholds are routinely assessed for each contact individually, a process that is uncomfortable and time-intensive, particularly with directional leads. The model could support this by providing imaging-based threshold estimates that narrow the clinical search space. Model performance to identify the directionality with lowest motor threshold was particularly strong (84%), which has a straightforward anatomical explanation: directional contacts are oriented at 120° intervals, and typically one faces posterolaterally toward the CSBT, producing a markedly lower threshold. Across-level prediction was less accurate (45%, p = 0.055), consistent with the CSBT running approximately parallel to the electrode axis. These results suggest the model is most informative where the clinical need is greatest: guiding directional steering of electric current to avoid motor side effects. Beyond manual programming, the model could be integrated into automated algorithms such as StimFit, which already uses side-effect probability as an optimization constraint.^20^

To support external evaluation and to provide a foundation for the integrations described above, the trained model is released as *CapsulePredictor*, a lightweight open-source Lead-DBS add-on (see *Suppl. Text 2*). Two considerations motivated the release of a self-contained model implementation. First, predictive models in DBS have so far rarely been made available, limiting external evaluation, comparison across cohorts, and methodological refinement. Second, the value of a probabilistic side-effect model lies primarily in its application to new patients; this requires that the full pipeline from electrode reconstruction through pathway activation modeling to probability estimation is accessible without custom implementation effort. In addition, integration into broader open-source DBS software such as StimFit^20^ and Lead-DBS^31^ is currently underway.

Several limitations should be considered when interpreting the results of this study.

The PLIC tract used in this study was constructed in MNI standard space and warped to individual patient space using nonlinear deformation fields. Patient-specific diffusion tractography can capture individual anatomical variability, and there is evidence that it may improve predictions for both therapeutic effects and PLIC-mediated side effects.^32,33^ At the same time, tractography results depend on acquisition protocol and reconstruction algorithm, factors that vary across clinical sites and can limit the reproducibility and wider deployment of models that rely on them.^34^ Furthermore, diffusion tractography produces inhomogeneous streamline densities that do not directly reflect underlying fiber densities,^35,36^ a property that requires careful consideration when used as input to pathway activation models. These considerations have motivated the development of curated anatomical pathway atlases to provide standardized tract representations independent of site-specific imaging protocols.^37-39^ In the present study, a normative PLIC tract was chosen to ensure reproducibility and to enable the open-source release of a model that does not require diffusion imaging data. Future work should evaluate whether patient-specific or hybrid approaches that combine normative and individual tract information could further improve prediction accuracy.

The empirical axon diameter distribution was derived from electron microscopy of the macaque pyramidal tract.^22^ While the macaque corticospinal tract shares key organizational features with the human tract, species differences in the absolute distribution of fiber diameters cannot be excluded. To our knowledge, direct histological characterization of the human corticospinal axon diameter distribution at the level of the STN is not yet available. The Firmin et al.^22^ distribution therefore represents the best available approximation, though future studies using human post-mortem tissue could refine these estimates.

While the anatomical back-projection of model coefficients revealed patterns consistent with known PLIC anatomy across both cohorts (*Fig. 4, Suppl. Fig. 5*), a general consideration applies: in physics-informed prediction frameworks, biophysically motivated features can yield good predictive performance even if the underlying biological assumptions are not entirely correct, because the statistical model can exploit any feature that covaries with the outcome regardless of whether it reflects the true causal mechanism. The anatomical mapping should therefore be considered as a tool for spatial orientation and plausibility assessment rather than a precise quantification of individual tract segment contributions.

The model does not capture somatotopic organization within the CSBT. Leg muscle contractions were rarely observed in our dataset, consistent with previous reports and the anatomical argument that leg-innervating fibers occupy the most posterolateral portion of the PLIC, positioned beyond the typical reach of DBS electric fields at standard stimulation amplitudes.^5,40^ The current dataset does not provide sufficient data to resolve within-tract somatotopy, and we deliberately refrain from making claims about somatotopic gradients.

The two cohorts were assessed under different clinical conditions (general anesthesia vs. awake, medication-ON state), which may affect muscle tone and the detectability of motor contractions. While the condition-specific interaction terms account for systematic differences, the individual contributions of clinical state, electrode type, and assessment protocol cannot be fully disentangled. The intraoperative cohort was collected retrospectively, which may introduce assessment variability. The slightly lower intraoperative model performance (AUC 0.95 vs. 0.97, COD 31% vs. 35%) may partly reflect this.

Dysarthria was deliberately excluded from the primary endpoint definition despite being commonly attributed to corticobulbar tract activation. However, the anatomical tract(s) involved in stimulation-induced dysarthria remain debated, and recent evidence suggests that corticobulbar as well as cerebellar projections are involved.^41^ In the postoperative cohort, dysarthria was assessed through general verbal interaction rather than a standardized speech test. When included alongside motor contractions in a sensitivity analysis, model performance remained comparable (*Suppl. Table 2*), which points towards a substantial corticobulbar involvement in stimulation-induced dysarthria in our cohort. However, by excluding dysarthria from our primary endpoint we opted for a more conservative primary endpoint to avoid conflating mechanistically distinct phenomena.

Finally, all data were collected at a single center using a standardized surgical and assessment protocol. Multi-center validation with different surgical teams, imaging protocols, and electrode types will be necessary to confirm the generalizability of the model for different clinical environments.

This study presents the first framework that bridges biophysically realistic pathway activation modeling and clinical outcome prediction for stimulation-induced motor contractions in DBS. The trained model is made publicly available through *CapsulePredictor* (see *Code Availability*), allowing patient-specific prediction with the aim of providing a foundation for personalized computational tools that support DBS treatment from surgical planning to postoperative programming.

## Methods

### 4.1 Study Design and Participants

Data were collected at Charité – Universitätsmedizin Berlin in two sequential study phases: an intraoperative phase conducted at the time of DBS surgery and a postoperative phase performed at least three months after DBS implantation. Intraoperative data were collected retrospectively from routinely acquired clinical records, whereas postoperative data were collected prospectively under a dedicated study protocol. The study was approved by the local ethics committee (EA4/230/21) and all participants provided written informed consent prior to postoperative assessments. Inclusion and exclusion criteria and a trial profile are provided in *Suppl Fig. 1*.

### 4.2 Surgical Procedure and Intraoperative Data Acquisition

All patients underwent STN-DBS implantation under general anesthesia following the standard surgical protocol of our center. On the day of surgery, a stereotactic frame (Leksell, Elekta Instrument AB, Stockholm, Sweden) was mounted to the patient’s skull, followed by a stereotactic CT that was co-registered to the preoperative MRI, allowing transformation of MRI-based target coordinates into the stereotactic reference frame (see *Section 4.4* for imaging details). STN target coordinates were determined by direct MRI-based targeting using Brainlab Elements stereotactic planning software (Brainlab SE, München, Germany).

Intraoperatively, two or three parallel test electrodes were advanced toward the STN target along the planned trajectory according to stereotactic frame coordinates. Electrical test stimulation was performed at two depth levels per trajectory, typically separated by 3 mm within the STN. Stimulation was delivered using the Neuro Omega system (Alpha Omega Engineering, Ziporit, Israel) in monopolar configuration with a pulse width of 60 µs and a frequency of 130 Hz. Stimulation amplitude was increased in 0.5 mA steps from 0 to a maximum of 7 mA or until motor contractions were observed.

Motor responses were assessed simultaneously by two independent modalities: visual inspection of the contralateral face, arm, and leg, and concurrent needle EMG recordings from the orbicularis oris, brachioradialis, thenar, and tibialis anterior muscles. The amplitudes at which motor contractions or EMG-motor unit activity were first observed were recorded as motor thresholds for each stimulation site, separately for clinical and EMG assessment. Following completion of test stimulation, the permanent DBS lead was implanted along the selected trajectory and height. The stimulation protocol was not modified for study purposes and reflects standard clinical practice at our center.

### 4.3 Postoperative Data Acquisition

Postoperative assessments were conducted prospectively at least three months after DBS implantation to allow for stabilization of the electrode-tissue interface and followed a similar stimulation protocol as described for the intraoperative phase. Assessments were performed in the medication-ON state to increase patient compliance and comfort. Each of the eight electrode contacts of the implanted directional DBS lead (Medtronic SenSight×, Medtronic, Minneapolis, MN, USA, *Fig. 3b*) was tested individually in monopolar configuration (monopolar review), in randomized order, with stimulation amplitude increased at a rate of 0.5 mA per 10 seconds up to a maximum of 7 mA or until motor contractions or intolerable side effects occurred (pulse width 60 µs, frequency 130 Hz). In case of transient side effects such as paraesthesias, the ramp was paused until symptoms resolved before continuing. Upon detection of motor contractions, the threshold was refined by stepwise amplitude reduction in 0.1 mA to the lowest amplitude producing sustained contractions. Motor responses were assessed by visual inspection of the contralateral face, arm, and leg. Parallel surface EMG recordings were obtained from the same muscle groups as in the intraoperative phase using the TMSI Porti system (TMSi, Oldenzaal, The Netherlands). Details of EMG acquisition and offline threshold analysis are provided in the *Suppl. Text 1*. Agreement between clinically observed and EMG-derived motor thresholds was assessed using Pearson correlation to confirm the consistency of the clinical endpoint across both assessment modalities. Clinically observed thresholds were used as the primary endpoint for all subsequent analyses.

### 4.4 Imaging and Electrode Reconstruction

Preoperative MRI was acquired as part of clinical routine using standardized high-resolution sequences on a Skyra 3T system (Siemens, Erlangen, Germany), including T1 (1 mm^3^ isovoxel), T2, and FGATIR sequences.^42^

Electrode localization followed different approaches for the two study phases, reflecting the different imaging data available at each study phase. Following the pipeline of Oxenford et al., intraoperative stimulation sites were localized based on stereotactic frame coordinates, which were registered to the preoperative MRI coordinate space for subsequent modeling.^30^ Postoperative DBS electrode locations were reconstructed from postoperative CT images co-registered to the preoperative MRI following the default Lead-DBS pipeline (v2.6 & v3.2).^43^ Results were visually validated to confirm accurate and complete preprocessing. To assess the internal consistency of both localization approaches, the Euclidean distance between intraoperative coordinate-based and postoperative CT-based electrode reconstructions was computed for each hemisphere.

For spatial normalization to MNI space, patient-specific nonlinear deformation fields were estimated by multimodal registration of preoperative MR sequences using the ANTs Symmetric Normalization algorithm followed by brain-shift correction as implemented in Lead-DBS.^43-45^ Normalization quality was assessed by visual inspection. Manual corrections were omitted to avoid introducing individual errors that could compromise reproducibility.

### 4.5 Anatomical Modeling of the Posterior Limb of the Internal Capsule

The PLIC was modeled as 1000 uniformly distributed streamlines, following the methodology described in Friedrich et al. and Sahin et al..^38,39^ In brief, tract boundaries at STN level were defined manually in MNI space based on anatomical literature,^46^ previously published white matter atlases,^37,47^ a high-resolution ex-vivo 7T MRI dataset,^48^ and volumetric atlas segmentation.^49,50^ Streamlines were seeded from the motor cortex to the cerebral peduncles and generated using the *CurveToBundle* module in 3D Slicer (https://github.com/netstim/SlicerNetstim.git). This approach ensures uniform spatial sampling across the tract cross-section. The tract is openly available as part of the FOCUS atlas^38^ and is additionally bundled with the *CapsulePredictor* software release described in *Section 4.10*.

To capture spatial heterogeneity of CSBT organization within the PLIC, the tract was subdivided into ten segments of equal fiber count, a granularity that maintains sufficient fiber counts per segment for robust activation estimates *(Fig. 1a)*. For each streamline, the mean transverse position at STN level was computed and projected onto the anteromedial-to-posterolateral axis. Fibers were then ranked along this axis and assigned to one of ten bins of equal fiber count. This segmentation was chosen to capture the known anteromedial-posterolateral gradient of corticospinal fiber density within the PLIC at this level^21^.

The patient-specific nonlinear deformation fields estimated in *Section 4.4* were applied to warp the normative PLIC segments from MNI space into individual patient space, preserving the ten-segment topology while accounting for individual anatomical variability. All subsequent biophysical simulations were performed on PLIC-segment representations in patient space.

### 4.6 Pathway Activation Modeling

Electric field and pathway activation modeling were performed using OSS-DBS v2.^31,51^ In detail, for each hemisphere, a patient-specific volume conductor model was constructed by assigning tissue-specific electrical conductivities to gray matter, white matter, and cerebrospinal fluid. Multispectral tissue segmentation was performed using the unified segmentation algorithm as implemented in SPM12^52^ and conductivity values were estimated based on the 4-Cole-Cole parametric model of Gabriel et al..^53^ The quasistatic formulation of Maxwell’s equations was solved using the Fourier Finite Element Method, with Dirichlet boundary conditions defined on the active electrode contacts and the outer surface of the computational domain representing case grounding.^8^ Non-active contacts were modelled as floating conductors. The resulting DBS-induced extracellular electric potential distribution was used as input to biophysical multi-compartment cable models based on the cable equation formulation of McIntyre et al.^9^, as implemented in OSS-DBS v2.

These cable models explicitly account for axon diameter, a key determinant of excitability in extracellular electric fields, with larger-diameter fibers exhibiting lower activation thresholds.^11,54^ Because the PLIC comprises axons spanning a wide range of diameters, this heterogeneity was explicitly modeled using the empirical corticospinal tract axon diameter distribution reported by Firmin et al..^22^ The distribution was binned in 1 µm steps from 2 to 10 µm, reflecting the lower bound of axon diameters for which reliable multi-compartment cable model parameters are available. Pathway activation modeling was performed separately for each diameter bin at amplitudes from 0.5 to 7.0 mA in 0.5 mA steps, matching the clinical stimulation protocol (*Sections 4.2 & 4.3*). The resulting activation estimates were weighted by the empirical fiber diameter distribution, yielding for each PLIC segment, stimulation site, and amplitude an activation profile representing the expected proportion of activated fibers for a biophysically plausible corticospinal axon population *(Fig. 1a)*.

### 4.7 Predictive Model

In pathway activation modeling, spatially adjacent tracts are necessarily co-activated by the electric field, producing correlated activation profiles. Collinear predictive features can, however, produce unstable parameter estimates in predictive models. PCA was therefore applied to extract orthogonal spatial activation patterns from the ten-dimensional segment-activation profiles *(Fig. 1b)*, eliminating collinearity among predictors and reducing the dimensionality of the feature space. PCA components were then retained as features for the subsequent predictive model.

Motor contraction probability was modeled using a GLM with a logistic link function. For each stimulation site, the clinical response at each tested amplitude step was encoded as a binary outcome variable (motor contraction present or absent). Amplitudes above the observed motor threshold were classified as contraction present, reflecting the physiological assumption that contractions persist at suprathreshold amplitudes. If stimulation was discontinued before reaching the maximum amplitude due to intolerable non-motor side effects, amplitude steps above the discontinuation threshold were excluded from the analysis.

Stimulation amplitude and the retained PCA scores were entered as main effects. Additionally, study condition (intraoperative vs. postoperative) was included as an interaction term with each main effect, allowing condition-specific coefficients while maintaining a shared model structure:

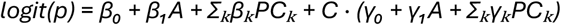

where *p* denotes the predicted motor contraction probability, *A* stimulation amplitude, *PC*_k_ the k-th retained principal component score, and *C* the binary study condition indicator (0 = intraoperative, 1 = postoperative). Model parameters were estimated by maximum likelihood on all available observations from both study phases.

The model thus estimates the probability of motor contractions for a given PLIC activation profile, stimulation amplitude, and study condition. To enable comparison with clinically observed motor thresholds, predicted thresholds were derived as the amplitude at which a 50% contraction probability was estimated, obtained by linear interpolation between adjacent amplitude steps.

### 4.8 Model Evaluation and Sensitivity Analysis

Model performance was evaluated using leave-one-patient-out cross-validation (LOOCV). Because the postoperative cohort was a sub-cohort of the intraoperative cohort, each patient’s data were excluded simultaneously from both conditions to prevent data leakage. Within each fold, PCA was recomputed on the training data, and the number of retained components was selected to maximize cross-validated threshold prediction performance (*Suppl Fig. 4*).

Classification performance was quantified by the area under the receiver operating characteristic curve (ROC-AUC, *Fig. 2a,c)*. To assess the discriminative contribution of PLIC activation independently of stimulation amplitude, classification performance was additionally evaluated at fixed amplitude levels within the clinically relevant range (2-5 mA, 1 mA steps). Threshold prediction performance was assessed using the coefficient of determination (COD) and mean absolute error (MAE) between predicted and observed motor thresholds *(Fig. 2b,d)*.

For the postoperative sub-cohort, the clinical utility of the model was additionally evaluated at the electrode level. For each directional DBS electrode, contacts were ranked by predicted motor threshold, and the correspondence between model-predicted and empirically observed rankings was assessed *(Fig. 3)*. Performance was quantified as the proportion of electrodes for which the model correctly identified the contact with the lowest (or highest) empirically observed threshold, separately for all contacts, directional contacts only, and directional contacts within the same electrode level. Chance-level performance was computed analytically compared to model performance using a one-sided exact Poisson-binomial test at an alpha level of 0.05.

To assess the sensitivity of the results to the choice of clinical endpoint and cohort composition, three complementary sensitivity analyses were performed. First, the model was re-estimated and evaluated using EMG-derived motor thresholds in place of clinically observed thresholds. Second, separate models were estimated on the intraoperative and postoperative cohorts independently to evaluate whether the results were driven by one cohort or contingent on the joint estimation across both study phases. In all analyses, the full modeling and cross-validation pipeline was repeated, and results were compared to the primary analysis. Finally, the effect of including dysarthria alongside motor contractions was assessed by re-estimating the model with an extended endpoint definition (*Suppl. Table 2*).

### 4.9 Anatomical Mapping and Coefficient Interpretation

To link model parameters to the underlying fiber anatomy, the GLM coefficients were projected from PCA space back onto the original ten PLIC segments. This was done separately for each condition (intra- and postoperative), by first computing effective regression coefficients that combine the respective main and condition-specific interaction effects for each principal component. These effective coefficients were then projected through the PCA loading matrix onto the ten segments, reversing the PCA transformation to express the model’s learned associations in the original anatomical space. The resulting weights reflect the relative contribution of each PLIC segment to the predicted motor contraction probability and were visualized as a function of segment position along the anteromedial-to-posterolateral axis *(Fig. 4a)*. To contextualize which PLIC regions were predominantly recruited across the dataset, mean fiber activation per segment was additionally computed at the condition-specific mean observed motor threshold amplitude (see *Section 2.2*) and visualized alongside the coefficient distribution *(Fig. 4b)*. Both the back-projected coefficients and the mean activation profile were mapped onto the PLIC streamlines, yielding complementary spatial representations of predicted motor contraction risk and actual fiber recruitment at the level of the STN.

### 4.10 Open-source implementation

To support external evaluation, reproducibility, and model comparison, the trained model was packaged as an open-source MATLAB add-on for Lead-DBS. This add-on, named *CapsulePredictor*, bundles the segmented PLIC, the GLM coefficients, and the PCA loadings described in *Sections 4.6 and 4.7*. It allows users to apply the trained model to new patients with stimulation settings of choice, requiring only a Lead-DBS electrode reconstruction as input. *CapsulePredictor* first invokes OSS-DBS to estimate PLIC activation and then applies the PCA loadings and GLM coefficients to predict motor contraction probabilities. Predictions and per-segment activations are stored within a BIDS-conformant directory inside the patient folder. A quick-start guide is provided in *Suppl. Text 2*.

### 4.11 Software Environment

Statistical modeling, cross-validation, and visualization were implemented in MATLAB R2024a (MathWorks, Natick, MA, USA). All analyses were implemented and executed by the authors without the use of artificial intelligence tools. Large language models (Claude Sonnet 4.6 and Claude Opus 4.6, Anthropic) were subsequently used for code refactoring and quality review of the analysis code without any consequential changes in the results or pipeline architecture itself. The manuscript was drafted by the authors; language editing was assisted by the same tools. All authors reviewed and take responsibility for the final content.

## Data availability

The individual-patient clinical and imaging data underlying this study contain sensitive health information and cannot be shared publicly under applicable data protection regulations (EU GDPR; ethics approval EA4/230/21). De-identified data sufficient to reproduce the main analyses are available from the corresponding author upon reasonable request, subject to approval by the local ethics committee and the execution of a data transfer agreement with Charité – Universitätsmedizin Berlin.

## Code availability

The biophysical modeling pipeline is based on the open-source tools OSS-DBS (https://github.com/SFB-ELAINE/OSS-DBS) and Lead-DBS (https://www.lead-dbs.org). The trained model is released as *CapsulePredictor*, a Lead-DBS add-on that allows users to apply the model described in this work to new patient data. The repository is available at https://github.com/JRoediger/CapsulePredictor and a versioned snapshot is archived at https://zenodo.org/records/20025108. All custom analysis code developed for this study, including the cross-validation pipeline, anatomical mapping, and visualization scripts, is available at https://github.com/JRoediger/CapsPAM with a versioned snapshot at https://zenodo.org/records/20038256. The repositories are currently under restricted access for peer review and will be made publicly available upon publication.

## Acknowledgments

This work was supported by the Deutsche Forschungsgemeinschaft (DFG, German Research Foundation) – Project-ID 424778381 – TRR 295 (A.A.K, G.H.S) and by Germany’s Excellence Strategy, NeuroCure Cluster of Excellence EXC 2049—390688087 (J.R., B.A., A.A.K.). I.A.S was funded by a scholarship from Einstein Center for Neurosciences. J.K.B. and M.S.T. are fellows of the BIH-Charité Junior Clinician Scientist Programme funded by Charité-Universitätsmedizin Berlin and Berlin Institute of Health (BIH). The funders played no role in study design, data collection, analysis and interpretation of data, or the writing of this manuscript.

## Author Contributions

J.R. conceptualized the study, developed the modeling and analysis pipeline, performed the statistical analysis, created the figures, and wrote the original draft. K.B. developed the OSS-DBS software, performed the biophysical pathway activation simulations, and contributed to the development of the computational methodology. A.P.K. and J.K.B. contributed to clinical data collection. I.A.S. constructed the normative PLIC tract. S.O. developed the intraoperative electrode localization pipeline. J.S., M.P., M.S.T., T.P., B.A.F., T.A.D. and A.A.K. contributed to the interpretation of results. G.H.S. performed the surgical procedures. A.A.K. supervised the study. All authors reviewed and edited the manuscript and approved the final version.

## References

1. Lozano AM, Lipsman N, Bergman H, et al. Deep brain stimulation: current challenges and future directions. Nat Rev Neurol 2019;15(3):148–160. DOI: 10.1038/s41582-018-0128-2.

2. Vedam-Mai V, Deisseroth K, Giordano J, et al. Proceedings of the Eighth Annual Deep Brain Stimulation Think Tank: Advances in Optogenetics, Ethical Issues Affecting DBS Research, Neuromodulatory Approaches for Depression, Adaptive Neurostimulation, and Emerging DBS Technologies. Front Hum Neurosci 2021;15:644593. DOI: 10.3389/fnhum.2021.644593.

3. Volkmann J, Moro E, Pahwa R. Basic algorithms for the programming of deep brain stimulation in Parkinson’s disease. Mov Disord 2006;21 Suppl 14:S284–9. DOI: 10.1002/mds.20961.

4. Hamani C, Saint-Cyr JA, Fraser J, Kaplitt M, Lozano AM. The subthalamic nucleus in the context of movement disorders. Brain 2004;127(Pt 1):4–20. DOI: 10.1093/brain/awh029.

5. Tommasi G, Krack P, Fraix V, et al. Pyramidal tract side effects induced by deep brain stimulation of the subthalamic nucleus. J Neurol Neurosurg Psychiatry 2008;79(7):813–9. DOI: 10.1136/jnnp.2007.117507.

6. Machado A, Rezai AR, Kopell BH, Gross RE, Sharan AD, Benabid AL. Deep brain stimulation for Parkinson’s disease: surgical technique and perioperative management. Mov Disord 2006;21 Suppl 14:S247–58. DOI: 10.1002/mds.20959.

7. Dembek TA, Reker P, Visser-Vandewalle V, et al. Directional DBS increases side-effect thresholds-A prospective, double-blind trial. Mov Disord 2017;32(10):1380– 1388. DOI: 10.1002/mds.27093.

8. Butson CR, McIntyre CC. Tissue and electrode capacitance reduce neural activation volumes during deep brain stimulation. Clin Neurophysiol 2005;116(10):2490–500. DOI: 10.1016/j.clinph.2005.06.023.

9. McIntyre CC, Richardson AG, Grill WM. Modeling the excitability of mammalian nerve fibers: influence of afterpotentials on the recovery cycle. J Neurophysiol 2002;87(2):995–1006. DOI: 10.1152/jn.00353.2001.

10. Gunalan K, Chaturvedi A, Howell B, et al. Creating and parameterizing patient-specific deep brain stimulation pathway-activation models using the hyperdirect pathway as an example. PLoS One 2017;12(4):e0176132. DOI: 10.1371/journal.pone.0176132.

11. Gunalan K, Howell B, McIntyre CC. Quantifying axonal responses in patient-specific models of subthalamic deep brain stimulation. Neuroimage 2018;172:263–277. DOI: 10.1016/j.neuroimage.2018.01.015.

12. Butson CR, Cooper SE, Henderson JM, McIntyre CC. Patient-specific analysis of the volume of tissue activated during deep brain stimulation. Neuroimage 2007;34(2):661–70. DOI: 10.1016/j.neuroimage.2006.09.034.

13. Chaturvedi A, Butson CR, Lempka SF, Cooper SE, McIntyre CC. Patient-specific models of deep brain stimulation: influence of field model complexity on neural activation predictions. Brain Stimul 2010;3(2):65–7. DOI: 10.1016/j.brs.2010.01.003.

14. Borgheai SB, Howell B, Isbaine F, et al. Evaluation of DBS computational modeling methodologies using in-vivo electrophysiology in Parkinson’s disease. Brain Stimul 2025;18(6):1996–2007. DOI: 10.1016/j.brs.2025.10.022.

15. Mahlknecht P, Akram H, Georgiev D, et al. Pyramidal tract activation due to subthalamic deep brain stimulation in Parkinson’s disease. Mov Disord 2017;32(8):1174–1182. DOI: 10.1002/mds.27042.

16. Dembek TA, Roediger J, Horn A, et al. Probabilistic sweet spots predict motor outcome for deep brain stimulation in Parkinson disease. Ann Neurol 2019;86(4):527–538. DOI: 10.1002/ana.25567.

17. Elias GJB, Boutet A, Joel SE, et al. Probabilistic Mapping of Deep Brain Stimulation: Insights from 15 Years of Therapy. Ann Neurol 2021;89(3):426–443. DOI: 10.1002/ana.25975.

18. Rajamani N, Friedrich H, Butenko K, et al. Deep brain stimulation of symptom-specific networks in Parkinson’s disease. Nat Commun 2024;15(1):4662. DOI: 10.1038/s41467-024-48731-1.

19. Roediger J, Dembek TA, Achtzehn J, et al. Automated deep brain stimulation programming based on electrode location: a randomised, crossover trial using a data-driven algorithm. Lancet Digit Health 2023;5(2):e59–e70. DOI: 10.1016/S2589-7500(22)00214-X.

20. Roediger J, Dembek TA, Wenzel G, Butenko K, Kuhn AA, Horn A. StimFit-A Data-Driven Algorithm for Automated Deep Brain Stimulation Programming. Mov Disord 2022;37(3):574–584. DOI: 10.1002/mds.28878.

21. Kretschmann HJ. Localisation of the corticospinal fibres in the internal capsule in man. J Anat 1988;160:219–25. (https://www.ncbi.nlm.nih.gov/pubmed/3253257).

22. Firmin L, Field P, Maier MA, et al. Axon diameters and conduction velocities in the macaque pyramidal tract. J Neurophysiol 2014;112(6):1229–40. DOI: 10.1152/jn.00720.2013.

23. Howell B, Isbaine F, Willie JT, et al. Image-based biophysical modeling predicts cortical potentials evoked with subthalamic deep brain stimulation. Brain Stimul 2021;14(3):549–563. DOI: 10.1016/j.brs.2021.03.009.

24. Butenko K, Li N, Neudorfer C, et al. Linking profiles of pathway activation with clinical motor improvements - A retrospective computational study. Neuroimage Clin 2022;36:103185. DOI: 10.1016/j.nicl.2022.103185.

25. Karniadakis GE, Kevrekidis IG, Lu L, Perdikaris P, Wang SF, Yang L. Physics-informed machine learning. Nat Rev Phys 2021;3(6):422–440. (In English). DOI: 10.1038/s42254-021-00314-5.

26. Butson CR, Maks CB, McIntyre CC. Sources and effects of electrode impedance during deep brain stimulation. Clin Neurophysiol 2006;117(2):447–54. DOI: 10.1016/j.clinph.2005.10.007.

27. Krakauer JW, Ghazanfar AA, Gomez-Marin A, MacIver MA, Poeppel D. Neuroscience Needs Behavior: Correcting a Reductionist Bias. Neuron 2017;93(3):480–490. DOI: 10.1016/j.neuron.2016.12.041.

28. Horn A, Fox MD. Opportunities of connectomic neuromodulation. Neuroimage 2020;221:117180. DOI: 10.1016/j.neuroimage.2020.117180.

29. Dembek TA, Baldermann JC, Petry-Schmelzer JN, et al. Sweetspot Mapping in Deep Brain Stimulation: Strengths and Limitations of Current Approaches. Neuromodulation 2022;25(6):877–887. DOI: 10.1111/ner.13356.

30. Oxenford S, Roediger J, Neudorfer C, et al. Lead-OR: A multimodal platform for deep brain stimulation surgery. Elife 2022;11. DOI: 10.7554/eLife.72929.

31. Neudorfer C, Butenko K, Oxenford S, et al. Lead-DBS v3.0: Mapping deep brain stimulation effects to local anatomy and global networks. Neuroimage 2023;268:119862. DOI: 10.1016/j.neuroimage.2023.119862.

32. Segura-Amil A, Nowacki A, Debove I, et al. Programming of subthalamic nucleus deep brain stimulation with hyperdirect pathway and corticospinal tract-guided parameter suggestions. Hum Brain Mapp 2023;44(12):4439–4451. DOI: 10.1002/hbm.26390.

33. Wang Q, Akram H, Muthuraman M, et al. Normative vs. patient-specific brain connectivity in deep brain stimulation. Neuroimage 2021;224:117307. DOI: 10.1016/j.neuroimage.2020.117307.

34. Maier-Hein KH, Neher PF, Houde JC, et al. The challenge of mapping the human connectome based on diffusion tractography. Nat Commun 2017;8(1):1349. DOI: 10.1038/s41467-017-01285-x.

35. Yeh CH, Jones DK, Liang X, Descoteaux M, Connelly A. Mapping Structural Connectivity Using Diffusion MRI: Challenges and Opportunities. J Magn Reson Imaging 2021;53(6):1666–1682. DOI: 10.1002/jmri.27188.

36. Zhang F, Daducci A, He Y, et al. Quantitative mapping of the brain’s structural connectivity using diffusion MRI tractography: A review. Neuroimage 2022;249:118870. DOI: 10.1016/j.neuroimage.2021.118870.

37. Petersen MV, Mlakar J, Haber SN, et al. Holographic Reconstruction of Axonal Pathways in the Human Brain. Neuron 2019;104(6):1056–1064 e3. DOI: 10.1016/j.neuron.2019.09.030.

38. Friedrich H, Sahin IA, Rajamani N, et al. A precise atlas of the human subcortex. bioRxiv 2026. DOI: 10.64898/2026.02.13.705755.

39. Sahin IA, Butenko K, Johnson KA, et al. Optimal Deep Brain Stimulation Locations for Gilles de la Tourette Syndrome. medRxiv 2026. DOI: 10.64898/2026.02.21.26346772.

40. Duerden EG, Finnis KW, Peters TM, Sadikot AF. Three-dimensional somatotopic organization and probabilistic mapping of motor responses from the human internal capsule. J Neurosurg 2011;114(6):1706–14. DOI: 10.3171/2011.1.JNS10136.

41. Jergas H, Petry-Schmelzer JN, Hannemann JH, et al. One side effect: two networks? Lateral and posteromedial stimulation spreads induce dysarthria in subthalamic deep brain stimulation for Parkinson’s disease. J Neurol Neurosurg Psychiatry 2025;96(3):280–286. DOI: 10.1136/jnnp-2024-333434.

42. Guttler C, Achtzehn J, Blomstedt P, et al. Toward a standard preoperative MRI protocol for functional neurosurgery. Imaging Neurosci (Camb) 2025;3. DOI: 10.1162/IMAG.a.52.

43. Horn A, Li N, Dembek TA, et al. Lead-DBS v2: Towards a comprehensive pipeline for deep brain stimulation imaging. Neuroimage 2019;184:293–316. DOI: 10.1016/j.neuroimage.2018.08.068.

44. Ewert S, Horn A, Finkel F, Li N, Kuhn AA, Herrington TM. Optimization and comparative evaluation of nonlinear deformation algorithms for atlas-based segmentation of DBS target nuclei. Neuroimage 2019;184:586–598. DOI: 10.1016/j.neuroimage.2018.09.061.

45. Avants BB, Epstein CL, Grossman M, Gee JC. Symmetric diffeomorphic image registration with cross-correlation: evaluating automated labeling of elderly and neurodegenerative brain. Med Image Anal 2008;12(1):26–41. DOI: 10.1016/j.media.2007.06.004.

46. Nieuwenhuys R, Voogd J, van Huijzen C. The Human Central Nervous System. Fourth Edition ed. Berlin Heidelberg New York: Springer-Verlag, 2008.

47. Meola A, Yeh FC, Fellows-Mayle W, Weed J, Fernandez-Miranda JC. Human Connectome-Based Tractographic Atlas of the Brainstem Connections and Surgical Approaches. Neurosurgery 2016;79(3):437–55. DOI: 10.1227/NEU.0000000000001224.

48. Edlow BL, Mareyam A, Horn A, et al. 7 Tesla MRI of the ex vivo human brain at 100 micron resolution. Sci Data 2019;6(1):244. DOI: 10.1038/s41597-019-0254-8.

49. Ewert S, Plettig P, Li N, et al. Toward defining deep brain stimulation targets in MNI space: A subcortical atlas based on multimodal MRI, histology and structural connectivity. Neuroimage 2018;170:271–282. DOI: 10.1016/j.neuroimage.2017.05.015.

50. Pijnenburg R, Scholtens LH, Ardesch DJ, de Lange SC, Wei Y, van den Heuvel MP. Myelo- and cytoarchitectonic microstructural and functional human cortical atlases reconstructed in common MRI space. Neuroimage 2021;239:118274. DOI: 10.1016/j.neuroimage.2021.118274.

51. Butenko K, Bahls C, Schroder M, Kohling R, van Rienen U. OSS-DBS: Open-source simulation platform for deep brain stimulation with a comprehensive automated modeling. PLoS Comput Biol 2020;16(7):e1008023. DOI: 10.1371/journal.pcbi.1008023.

52. Ashburner J, Friston KJ. Unified segmentation. Neuroimage 2005;26(3):839–51. DOI: 10.1016/j.neuroimage.2005.02.018.

53. Gabriel S, Lau RW, Gabriel C. The dielectric properties of biological tissues: III. Parametric models for the dielectric spectrum of tissues. Phys Med Biol 1996;41(11):2271–93. DOI: 10.1088/0031-9155/41/11/003.

54. McNeal DR. Analysis of a model for excitation of myelinated nerve. IEEE Trans Biomed Eng 1976;23(4):329–37. DOI: 10.1109/tbme.1976.324593.

