## Supplementary Material for "Biophysical modeling of corticospinal tract activation predicts motor contractions in subthalamic deep brain stimulation"

### Supplementary Figures

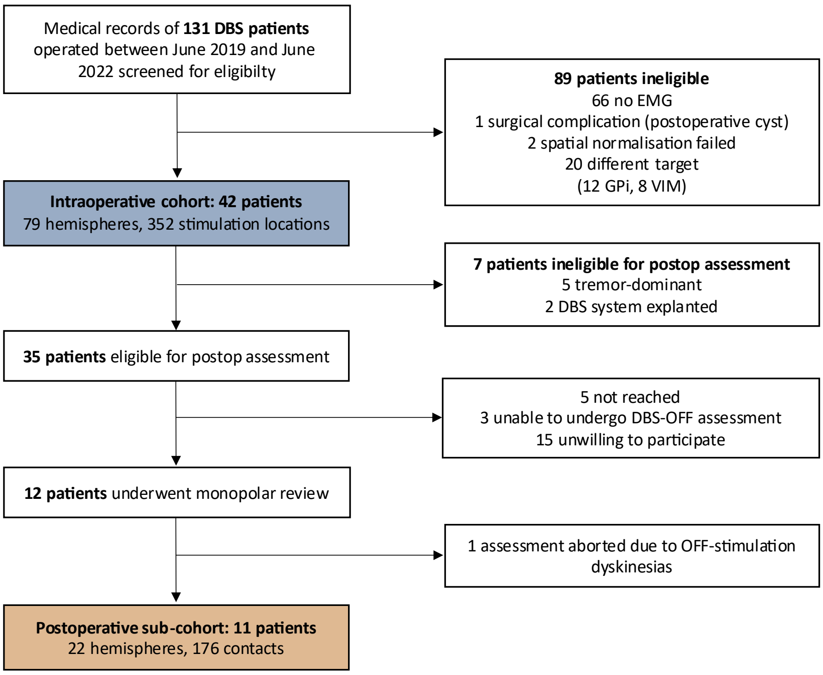


**Supplementary Figure 1 | Trial profile.** Flow diagram of participant enrollment across the two sequential study phases: the retrospectively collected intraoperative cohort (blue) and the prospectively collected postoperative sub-cohort (orange), comprising patients who underwent DBS implantation between June 2019 and June 2022. Intraoperative EMG acquisition in clinical routine was performed only when equipment and personnel were available in the operating theatre, accounting for the majority of intraoperative exclusions. Of the 84 hemispheres in the 42 included patients, five were excluded due to unavailability of intraoperative EMG, yielding 79 hemispheres for analysis. Tremor-dominant disease phenotype was an exclusion criterion for postoperative assessment, as persistent tremor may compromise motor contraction detection, particularly via EMG. DBS-OFF denotes the baseline state with deep brain stimulation switched off, as required for monopolar review.

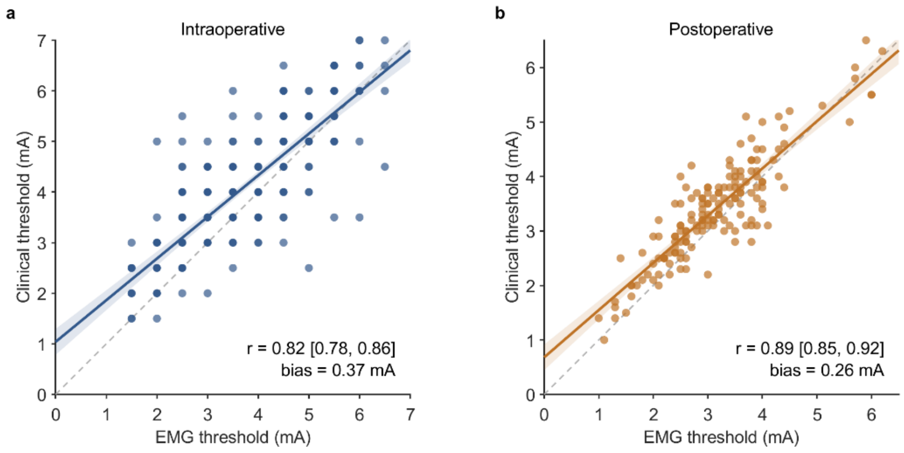


**Supplementary Figure 2 | Agreement between clinical and EMG-derived motor contraction thresholds.** Scatter plots comparing visually observed and EMG-derived motor thresholds for the intraoperative (a) and postoperative (b) cohorts, with each point representing one stimulation site (intraoperative) or contact (postoperative). The dashed gray line indicates identity (y = x); the solid line shows the linear regression fit with 95% confidence band. Inset statistics report Pearson's correlation coefficient (r) with 95% confidence interval and the mean bias (clinical - EMG).

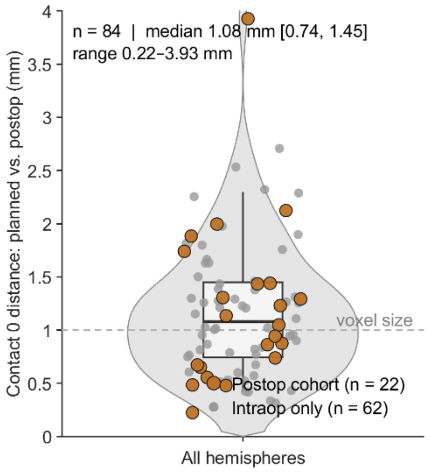


**Supplementary Figure 3 | Agreement between intraoperative and postoperative electrode localization.** Euclidean distance between the planned electrode tip position (based on intraoperative stereotactic coordinates) and the center of the ventral contact (level 0) of the implanted DBS electrode as reconstructed from postoperative CT, computed in patient native space for each hemisphere of the intraoperative cohort (n = 84 hemispheres). Hemispheres belonging to patients of the postoperative sub-cohort (n = 22) are highlighted in orange; remaining hemispheres (n = 62) are shown in gray. The violin shows the kernel density of distances across all hemispheres; the embedded boxplot indicates median and IQR. The dashed horizontal line marks the 1 mm voxel resolution of the preoperative MRI for reference. Summary statistics (n, median [IQR], range) across all hemispheres are reported as an inset.

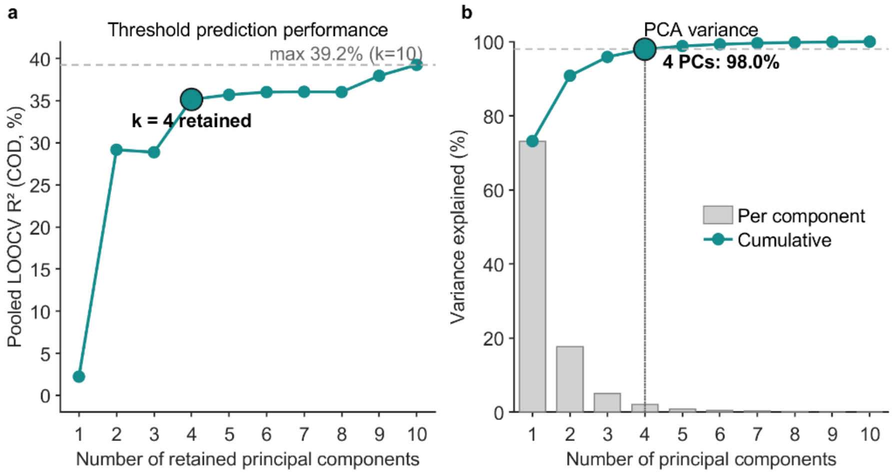


**Supplementary Figure 4 | Selection of the number of retained principal components.** Cross-validated threshold prediction performance (coefficient of determination, $R^2$) as a function of the number of retained principal components (k = 1 to 10). For each value of k, the full leave-one-patient-out cross-validation pipeline was run, and $R^2$ was computed from the pooled out-of-fold threshold predictions across both cohorts. Prediction performance increased substantially up to k = 4 and then reached a plateau; four components (highlighted) were therefore retained. A small further increase at k = 9-10 was observed, but these components individually contributed only 0.12% and 0.08% of the input variance, respectively (panel b), and were therefore considered likely to reflect unstable, low-variance features. (**b**) Variance of the ten-dimensional PLIC segment-activation profiles explained by each principal component (gray bars) and cumulatively (line). The first four components together account for 98% of the total variance.

**Supplementary Figure 5 | Anatomical mapping of model coefficients and fiber activation for the intraoperative cohort.** Analogous to main Figure 4, showing the back-projected GLM coefficients and mean fiber activation across the ten PLIC segments for the intraoperative cohort. Segments are ordered from anteromedial (I) to posterolateral (X). (**a**) Back-projected GLM coefficients reflecting the relative contribution of each segment to predicted motor contraction probability in the intraoperative condition (logit scale). Positive values indicate that activation of a given segment increases the predicted probability of motor contractions. Error bars denote 95% confidence intervals derived via the delta method. (**b**) Mean fiber activation (%) per PLIC segment at the intraoperative mean observed motor threshold amplitude (3.9 mA). Error bars denote 95% confidence intervals of the mean. The contrasting gradients across panels illustrate that the segments most predictive of motor contractions (posterolateral, panel a) are those least activated at typical therapeutic stimulation amplitudes (panel b), reproducing the pattern observed in the postoperative cohort.

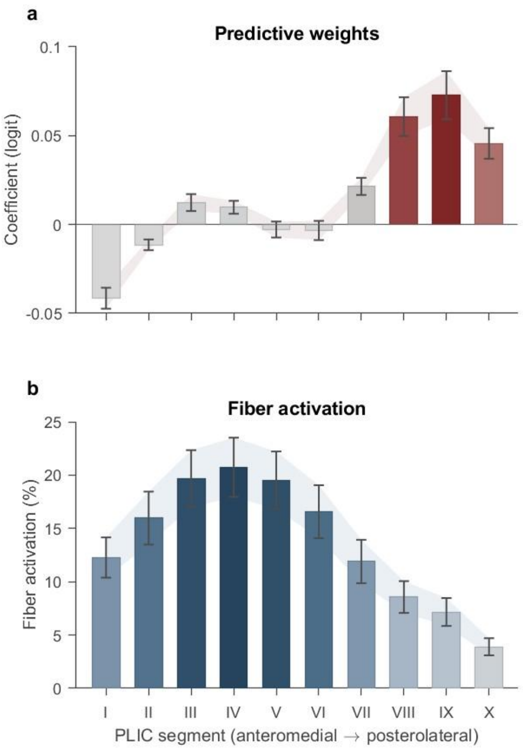


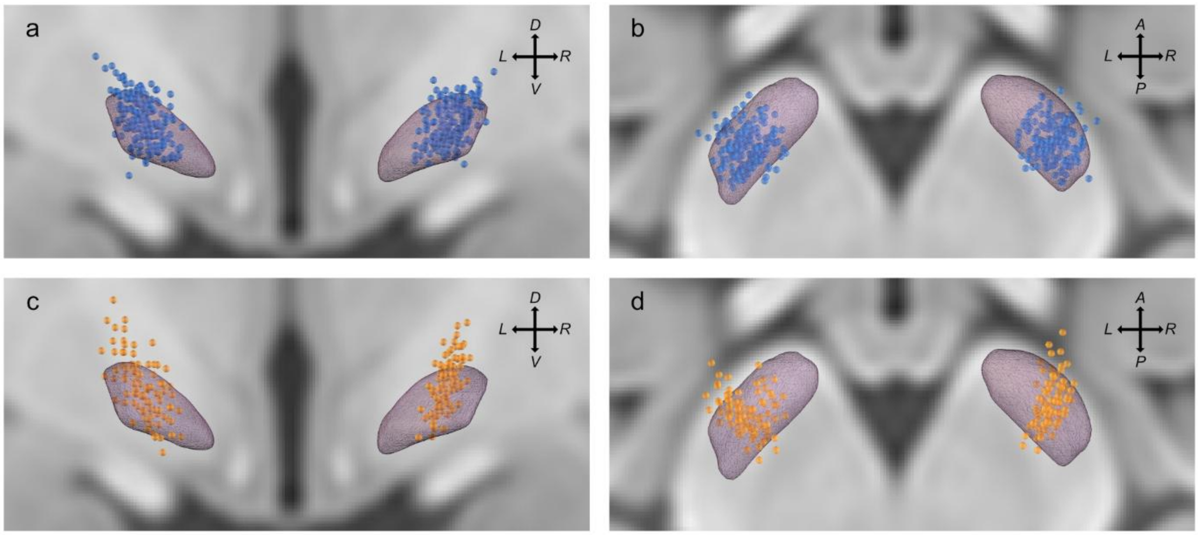


**Supplementary Figure 6 | Anatomical distribution of tested stimulation locations in MNI space.** Spatial distribution of all tested stimulation locations of the intraoperative (**a**, **b**; n = 352 sites, blue) and postoperative (**c**, **d**; n = 176 contacts, orange) cohorts, shown from a coronal (**a**, **c**) and axial (**b**, **d**) perspective in standard MNI-space. The subthalamic nucleus (DISTAL atlas) is shown in purple. D: dorsal; V: ventral; A: anterior; P: posterior; L: left; R: right.

### Supplementary Tables

| term | estimate | std_error | z_value | p_value | ci_lower | ci_upper |
|---|---|---|---|---|---|---|
| **Intercept** | -5.206 | 0.191 | -27.24 | <0.001 | -5.581 | -4.831 |
| **Amplitude** | 1.294 | 0.043 | 30.28 | <0.001 | 1.211 | 1.378 |
| **PC1** | 0.012 | 0.002 | 7.56 | <0.001 | 0.009 | 0.015 |
| **PC2** | -0.074 | 0.004 | -18.02 | <0.001 | -0.082 | -0.066 |
| **PC3** | 0.043 | 0.007 | 5.90 | <0.001 | 0.029 | 0.058 |
| **PC4** | 0.079 | 0.008 | 9.81 | <0.001 | 0.063 | 0.095 |
| **Condition (post vs intraop)** | 2.340 | 0.563 | 4.16 | <0.001 | 1.236 | 3.444 |
| **Amplitude × Condition** | 0.417 | 0.115 | 3.63 | <0.001 | 0.192 | 0.642 |
| **PC1 × Condition** | 0.070 | 0.009 | 7.69 | <0.001 | 0.052 | 0.088 |
| **PC2 × Condition** | -0.140 | 0.022 | -6.29 | <0.001 | -0.183 | -0.096 |
| **PC3 × Condition** | 0.155 | 0.030 | 5.19 | <0.001 | 0.097 | 0.214 |
| **PC4 × Condition** | 0.123 | 0.023 | 5.38 | <0.001 | 0.078 | 0.168 |

**Supplementary Table 1 | Parameter estimates of the joint generalized linear model.** Full fixed-effect coefficient table for the generalized linear model with logistic link function, estimated on all 7,261 binary observations across both cohorts. The reference category for the condition indicator is intraoperative, interaction terms therefore reflect the additional effect specific to the postoperative cohort. All terms reached statistical significance at $p < 0.001$. PC: principal component; CI: confidence interval.

| Model_variant | AUC | | COD | | MAE | |
|---|---|---|---|---|---|---|
| | intraop | postop | intraop | postop | intraop | postop |
| **Primary model** | **0.946** | **0.970** | **30.9** | **35.3** | **0.836** | **0.592** |
| **EMG-derived thresholds** | 0.930 | 0.963 | 24.6 | 32.8 | 0.920 | 0.666 |
| **Intraop-only model** | 0.946 | - | 30.8 | - | 0.835 | - |
| **Postop-only model** | - | 0.971 | - | 39.1 | - | 0.570 |
| **Dysarthria-extended endpoint** | - | 0.969 | - | 32.6 | - | 0.577 |

**Supplementary Table 2 | Model performance across sensitivity analyses.** Performance metrics of the primary model and four alternative specifications: (i) using EMG-derived rather than clinically observed motor contraction thresholds as the outcome definition; (ii) fitting the model separately on the intraoperative cohort; (iii) fitting the model separately on the postoperative cohort; and (iv) extending the endpoint to include stimulation-induced dysarthria alongside motor contractions (postoperative cohort only; defined as the occurrence of either motor contractions or dysarthria, whichever occurred at the lower amplitude). Dashes denote non-applicable combinations for single-cohort models. All performance metrics reached statistical significance at $p < 0.001$. AUC: area under the receiver operating characteristic curve; COD: coefficient of determination; MAE: mean absolute error of predicted motor threshold.

### Supplementary Text

#### Supplementary Text 1: EMG acquisition and threshold analysis

Intraoperatively, needle EMG was recorded bilaterally from four muscles: orbicularis oris, brachioradialis, thenar, and tibialis anterior. Two disposable subdermal needle electrodes per muscle (Neurodart, Spes Medica S.r.l., Genova, Italy) were inserted into the muscle belly at an interelectrode distance of 1-2 cm. A common ground/reference was placed over the anterior superior iliac spine. Signals were acquired through the Neuro Omega system (Alpha Omega Engineering, Ziporit, Israel) in bipolar configuration at a sampling rate of 11 kHz with an analog hardware bandpass filter of 0.075 Hz - 3.5 kHz, an online digital high-pass filter at 12 Hz, and an amplifier gain of 55. During each test stimulation, EMG signals were monitored live on the Neuro Omega display by a second rater independent of the clinician controlling the stimulation. For each of the four recorded muscles, the threshold was defined as the lowest stimulation amplitude at which a reproducible interference pattern emerged above baseline activity. Thresholds were determined per muscle in real time.

Postoperatively, surface EMG was recorded from the same four muscle groups, with orbicularis oculi replacing orbicularis oris to accommodate awake recording conditions. Disposable pre-gelled Ag/AgCl snap surface electrodes (Kendall™ H124SG, Cardinal Health, Dublin, OH, USA; 24 mm outer diameter, 10 mm Ag/AgCl sensor) were placed on the muscle bellies at an interelectrode distance of 1-2 cm, with the ground/reference electrode positioned over the anterior superior iliac spine. Signals were recorded using a TMSi Porti system (TMSi, Oldenzaal, The Netherlands) in bipolar configuration at a sampling rate of 2048 Hz, with a digital anti-aliasing low-pass filter at 0.27 × sampling frequency (~553 Hz) implemented in the analog-to-digital converter; no online high-pass or notch filter was applied. An additional channel was placed over the implanted pulse generator to capture the stimulation artefact, enabling temporal synchronization of the EMG recording with the stimulation amplitude ramp. EMG thresholds were determined offline in a semi-automated procedure by a single rater, blinded to the order of electrode contacts. Recordings were high-pass filtered offline at 1 Hz. The stimulation amplitude was reconstructed from the pulse-generator reference channel via rectification and Gaussian smoothing of the signal envelope, calibrated against the maximum tested amplitude documented per contact and verified and manually corrected by visual inspection. For each contact trial and each muscle, the rectified first derivative of the EMG signal was computed and displayed as a function of the reconstructed stimulation amplitude. Threshold selection proceeded in two stages. In the first stage, the rater assessed overall channel usability across all contacts of a session; channels with persistent noise, electrode loosening, or other systematic artefacts were excluded from threshold determination. In the second stage, for each usable channel, the rater inspected each contact trial individually and marked the threshold as the lowest stimulation amplitude at which the signal departed from baseline in a sustained, monotone-increasing manner. If no reproducible response was observed up to the maximum tested amplitude, the threshold was coded as "not reached" and the site was excluded from subsequent agreement and sensitivity analyses.

In both settings, thresholds were determined separately per muscle and the lowest threshold across muscles was taken as the EMG-derived motor contraction threshold for each stimulation site. This reflects the clinical definition of the motor contraction threshold as the amplitude at which contraction first appears in any contralateral muscle group. Operator judgement was the primary mechanism for artefact exclusion; no automated rejection was performed.

#### Supplementary Text 2: CapsulePredictor

*CapsulePredictor* is an open-source MATLAB add-on for Lead-DBS that applies the trained model described in this work to new patient data. The repository, including full documentation, is available at https://github.com/JRoediger/CapsulePredictor.

*Prerequisites. CapsulePredictor* requires a recent installation of Lead-DBS v3 (develop branch) and MATLAB R2024a or newer. The OSS-DBS environment is installed automatically on first use.

*Installation*. The repository is cloned to any local path. After changing into the cloned folder in MATLAB, the command setupPath adds *CapsulePredictor* to the MATLAB path for the current session. On first launch, the bundled PLIC_Sahin2026 tract is automatically copied into the Lead-DBS connectome directory so that OSS-DBS can resolve it during pathway activation modeling. No further setup is required.

*Workflow*. The add-on is invoked by capsulePredictor(), which opens a folder dialog, or by capsulePredictor(patientDir) with the patient path passed directly. The patient input is a Lead-DBS BIDS subject derivative folder (derivatives/leaddbs/sub-XXX/) with a completed postoperative electrode reconstruction. Both four-contact ring electrodes and eight-contact directional electrodes are supported. After patient selection, a graphical interface presents an editable stimulation table; multiple stimulation settings can be entered at once for batch processing. Stimulation settings are defined by amplitude, pulse width, and the percentage distribution of electric current across the pulse generator case and electrode contacts. Charge balance (anodic and cathodic currents summing to zero) is required for the pipeline to run. Clicking *Run* executes the OSS-DBS pathway activation simulation and the GLM prediction sequentially for each row whose prediction has not yet been computed, with a runtime of approximately 10–20 minutes per row on a typical workstation. Results are written to <patient>/stimulations/predictions/MotorContractions_Roediger2026/StimTable.mat and persisted after every completed row, so an interrupted batch can be resumed without loss of work. Each row stores the predicted motor contraction probability, the corresponding 95% confidence interval on the probability scale, and the ten axon-diameter–weighted segment activations (PLIC_I to PLIC_X) used as input to the model.

*Scope and limitations*. *CapsulePredictor* is intended exclusively for research purposes and is not a medical device; it must not be used to inform clinical decisions, treatment selection, DBS programming, or surgical planning. The current release supports predictions only for postoperative electrodes reconstructed with Lead-DBS; an extension to intraoperative electrodes based on surgical planning coordinates is underway. The model was trained exclusively on monopolar stimulation at a pulse width of 60 µs and a case fraction of +100% (anode on the implanted pulse generator); predictions outside this regime are extrapolations and may not be accurate. *CapsulePredictor* is distributed under the PolyForm Noncommercial 1.0.0 license; the full disclaimer and license terms are provided with the software repository.
